# Duration of protection of BNT162b2 and mRNA-1273 COVID-19 vaccines against symptomatic SARS-CoV-2 Omicron infection in Qatar

**DOI:** 10.1101/2022.02.07.22270568

**Authors:** Hiam Chemaitelly, Houssein H. Ayoub, Sawsan AlMukdad, Patrick Tang, Mohammad R. Hasan, Hadi M. Yassine, Hebah A. Al Khatib, Maria K. Smatti, Peter Coyle, Zaina Al Kanaani, Einas Al Kuwari, Andrew Jeremijenko, Anvar Hassan Kaleeckal, Ali Nizar Latif, Riyazuddin Mohammad Shaik, Hanan F. Abdul Rahim, Gheyath K. Nasrallah, Mohamed Ghaith Al Kuwari, Adeel A. Butt, Hamad Eid Al Romaihi, Mohamed H. Al-Thani, Abdullatif Al Khal, Roberto Bertollini, Laith J. Abu-Raddad

## Abstract

**BACKGROUND:** Qatar has been experiencing a large SARS-CoV-2 Omicron (B.1.1.529) wave that started on December 19, 2021. We assessed duration of protection of BNT162b2 (Pfizer-BioNTech) and mRNA-1273 (Moderna) COVID-19 vaccines after second dose and after third/booster dose against symptomatic Omicron infection and against COVID-19 hospitalization and death, between December 23, 2021 and February 2, 2022.

**METHODS:** Vaccine effectiveness was estimated using the test-negative, case-control study design, applying the same methodology used earlier to assess waning of BNT162b2 and mRNA-1273 effectiveness in the same population during earlier infection waves.

**RESULTS:** BNT162b2 effectiveness against symptomatic Omicron infection was highest at 61.9% (95% CI: 49.9-71.1%) in the first month after the second dose, but then gradually declined and was at 10% or less starting from the 5^th^ month after the second dose. After the booster, effectiveness rapidly rebounded to peak at about 55% between 2-5 weeks after the booster, but then started to decline again thereafter. Effectiveness against severe, critical, or fatal COVID-19 was maintained at >70% after the second dose and at >90% after the booster with no evidence for declining effectiveness over time. mRNA-1273 effectiveness against symptomatic Omicron infection was highest at 44.8% (95% CI: 16.0-63.8%) in the first three months after the second dose, before gradually declining to negligible levels thereafter. After the booster, effectiveness rapidly rebounded to peak at about 55% between 2-5 weeks after the booster, but then declined again thereafter. Effectiveness against severe, critical, or fatal COVID-19 was high at >60% after the second dose and at >80% after the booster, but the confidence intervals were wide owing to the small number of cases.

**CONCLUSIONS:** BNT162b2 and mRNA-1273 vaccines show a similar level and pattern of protection against symptomatic Omicron infection. Protection against Omicron is lower than that against Alpha, Beta, and Delta variants, and wanes more rapidly than against earlier variants after the second and booster doses. Meanwhile, protection against hospitalization and death appears robust and durable after both the second and booster doses.

## Introduction

Qatar has been experiencing a large severe acute respiratory syndrome coronavirus 2 (SARS-CoV-2) Omicron (B.1.1.529)^1^ wave that started on December 19, 2021 and peaked in mid-January, 2022.^2,3^ We assessed the duration of protection of BNT162b2 (Pfizer-BioNTech) and mRNA-1273 (Moderna) coronavirus disease 2019 (COVID-19) vaccines after second dose and after third/booster dose against symptomatic Omicron infection and against COVID-19 hospitalization and death, between December 23, 2021 and February 2, 2022.

## Methods

### Study population, data sources, and study design

This study was conducted in the resident population of Qatar, applying the same methodology that was used to assess waning of the BNT162b2^4^ and mRNA-1273^5^ COVID-19 vaccine effectiveness in the same population during earlier infection waves. A detailed description of this methodology can be found in Chemaitelly *et al*.^4^

COVID-19 laboratory testing, vaccination, clinical infection data, and related demographic details were extracted from the national, federated SARS-CoV-2 databases that include all polymerase chain reaction (PCR) testing, COVID-19 vaccinations, and COVID-19 hospitalizations and deaths in Qatar since the start of the pandemic.

Every PCR test conducted in Qatar is classified on the basis of symptoms and the reason for testing (clinical symptoms, contact tracing, surveys or random testing campaigns, individual requests, routine healthcare testing, pre-travel, at port of entry, or other). Qatar has unusually young, diverse demographics, in that only 9% of its residents are ≥50 years of age, and 89% are expatriates from over 150 countries.^6,7^ Nearly all individuals were vaccinated in Qatar, but if vaccinated elsewhere, those vaccinations were still recorded in the health system at the port of entry upon return to Qatar.

By February 2, 2022 (end of study), 1,301,203 individuals received ≥2 BNT162b2 doses in Qatar, of whom 301,059 received a booster dose. Meanwhile, 891,810 individuals received ≥2 mRNA-1273 doses, of whom 105,558 received a booster dose. The median dates of first, second, and third doses were May 3, 2021, May 24, 2021, and December 17, 2021 for BNT162b2, and May 28, 2021, June 27, 2021, and January 4, 2022 for mRNA-1273, respectively. The median time between the first and second dose was 21 days (interquartile range (IQR), 21-22 days) for BNT162b2 and 28 days (IQR, 28-30 days) for mRNA-1273, while that between second and booster dose was 250 days (IQR, 232-272 days) for BNT162b2 and 233 days (IQR, 212-254 days) for mRNA-1273.

Vaccine effectiveness during the large SARS-CoV-2 Omicron wave in Qatar, between December 23, 2021 and February 2, 2022, was estimated using the test-negative, case-control study design, a standard design for assessing vaccine effectiveness.^8-16^ Cases (PCR-positive persons) and controls (PCR-negative persons) were exact-matched two-to-one by sex, 10-year age group, nationality, and calendar week of PCR test to estimate effectiveness against symptomatic SARS-CoV-2 Omicron infection. Symptomatic SARS-CoV-2 Omicron infection was defined as a PCR-positive swab collected during the Omicron wave because of clinical suspicion due to presence of symptoms compatible with a respiratory tract infection.

The two-to-one matching ratio was done to improve the statistical precision of the estimates. This specific matching ratio was dictated by the higher number of PCR-positive tests than PCR-negative tests during the large Omicron wave (Figure S1). Matching was performed to control for known differences in the risk of exposure to SARS-CoV-2 infection in Qatar.^7,17-20^

Vaccine effectiveness was also estimated against any severe, critical, or fatal COVID-19 using the same methodology. Here, however, the matching ratio was one-to-five to improve the statistical precision given the relatively small number of severe, critical, and fatal COVID-19 cases during the Omicron wave.

Only the first PCR-positive test during the study was included for each case, whereas all PCR-negative tests during the study were included for each control. Controls included individuals with no record of a PCR-positive test during the study period. Only PCR tests conducted for clinical suspicion; that is due to presence of symptoms compatible with a respiratory tract infection, were included in the analysis. All persons who received a vaccine other than BNT162b2 or mRNA-1273, or who received mixed vaccines, were excluded. These inclusion and exclusion criteria were implemented to minimize different types of potential bias based on earlier analyses in the same population.^4,5^ Every control that met the inclusion criteria and that could be matched to a case was included in the analysis. Both PCR-test outcomes and vaccination status were ascertained at the time of the PCR test.

Classification of COVID-19 case severity (acute-care hospitalizations),^21^ criticality (ICU hospitalizations),^21^ and fatality^22^ followed World Health Organization (WHO) guidelines, and assessments were made by trained medical personnel using individual chart reviews (Section S1). Each person who had a positive PCR test result and COVID-19 hospital admission was subject to an infection severity assessment every three days until discharge or death, regardless of the length of the hospital stay or the time between the PCR-positive test and the final disease outcome. Individuals who progressed to severe,^21^ critical,^21^ or fatal^22^ COVID-19 between the PCR-positive test result and the end of the study were classified based on their worst outcome, starting with death, followed by critical disease, and then severe disease.

### Laboratory methods

Details of laboratory methods for real-time reverse-transcription PCR (RT-qPCR) testing and multiplex RT-qPCR variant screening are found in Section S2. All PCR testing was conducted at the Hamad Medical Corporation Central Laboratory or at Sidra Medicine Laboratory, following standardized protocols.

### Oversight

Hamad Medical Corporation and Weill Cornell Medicine-Qatar Institutional Review Boards approved this retrospective study with waiver of informed consent. The study was reported following the STROBE guidelines. The STROBE checklist is found in Table S1.

### Statistical analysis

All records of PCR testing in Qatar during the study were examined, but only samples of matched cases and controls that underwent PCR testing for clinical suspicion were included in the analysis. Demographic characteristics of study samples were described using frequency distributions and measures of central tendency. Study groups were compared using standardized mean differences (SMDs), defined as the difference in the mean of a covariate between groups divided by the pooled standard deviation. SMD <0.1 indicated adequate matching.^23^

The odds ratio, comparing odds of vaccination among cases versus controls, and its associated 95% confidence interval (CI) were derived using conditional logistic regression, that is factoring the matching in the study design. This matching and analysis approach aims to minimize potential bias due to variation in epidemic phase,^8,24^ gradual roll-out of vaccination during the study,^8,24^ or other confounders.^25,26^ 95% CIs were not adjusted for multiplicity and thus should not be used to infer definitive differences between different groups. Interactions were not investigated. Vaccine effectiveness at different time points and its associated 95% CI were calculated by applying the following equation:^8,9^

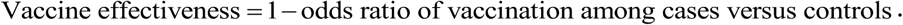

In each analysis for a specific time-since-vaccination stratum, we included only those vaccinated in that specific time-since-vaccination stratum and those unvaccinated (our reference group). Only matched pairs of PCR-positive and PCR-negative persons, in which members of the pair were either unvaccinated or fell within each time-since-vaccination stratum were included in the corresponding vaccine effectiveness estimate. Thus, the number of cases (and controls) varied across time-since-vaccination analyses. Effectiveness after the second dose was estimated by one or more months where one month was defined as 30 days. Effectiveness after the third/booster dose was estimated by one or more weeks where one week was defined as 7 days.

## Results

Figure S1 shows the process of selecting the study populations and Table S2 describes their demographic characteristics. The study was based on the total population of Qatar and thus the study population is broadly representative of the diverse, by national background, but young and predominantly male, total population of Qatar (Table S3).

BNT162b2 effectiveness against symptomatic Omicron infection was highest at 61.9% (95% CI: 49.9-71.1%) in the first month after the second dose, but then gradually declined and was at 10% or less starting from the 5^th^ month after the second dose (Figure 1 and Table 1). After the booster, effectiveness rapidly rebounded to peak at about 55% between 2-5 weeks after the booster, but then started to decline again thereafter. Effectiveness against severe, critical, or fatal COVID-19 was maintained at >70% after the second dose and at >90% after the booster with no evidence for declining effectiveness over time (Figure 1 and Table 2).

**Figure 1.**
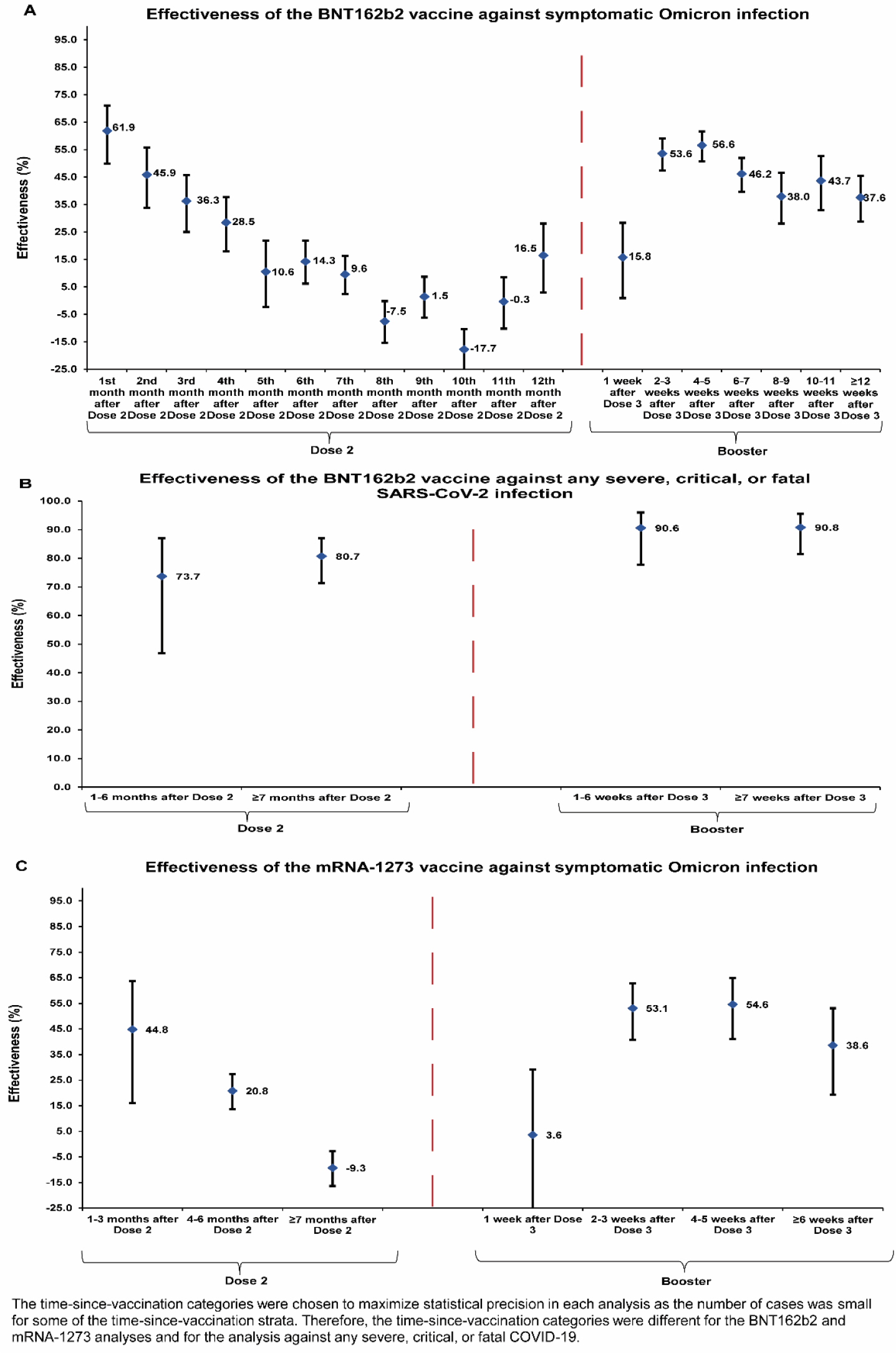
Effectiveness of the BNT162b2 vaccine against A) symptomatic SARS-CoV-2 Omicron infection and B) severe, critical, or fatal COVID-19 due to Omicron infection. C) Effectiveness of the mRNA-1273 vaccine against symptomatic SARS-CoV-2 Omicron infection. Data are presented as effectiveness point estimates. Error bars indicate the corresponding 95% confidence intervals.

**Table 1.** Effectiveness of the BNT162b2 and mRNA-1273 vaccines against symptomatic SARS-CoV-2 Omicron infection^*^.

| Sub-studies<sup>†</sup> | Cases<sup>‡</sup><br>(PCR-positive) |  | Controls<sup>‡</sup><br>(PCR-negative) |  | Effectiveness in %<br>(95% CI)<sup>§</sup> |
| --- | --- | --- | --- | --- | --- |
|  | Vaccinated | Unvaccinated | Vaccinated | Unvaccinated |  |
| **BNT162b2** |  |  |  |  |  |
| **Dose 1** |  |  |  |  |  |
| 0-13 days after Dose 1 | 60 | 12,866 | 37 | 7,781 | -7.2 (-40.3 to 38.6) |
| ≥14 days after Dose 1 and no Dose 2 | 155 | 12,889 | 127 | 7,784 | 26.1 (5.8 to 42.0) |
| **Dose 2** |  |  |  |  |  |
| 1<sup>st</sup> month after Dose 2 and no Dose 3 | 90 | 12,952 | 134 | 7,777 | 61.9 (49.9 to 71.1) |
| 2<sup>nd</sup> month after Dose 2 and no Dose 3 | 207 | 13,009 | 209 | 7,802 | 45.9 (33.8 to 55.8) |
| 3<sup>rd</sup> month after Dose 2 and no Dose 3 | 340 | 13,075 | 302 | 7,797 | 36.3 (25.1 to 45.8) |
| 4<sup>th</sup> month after Dose 2 and no Dose 3 | 524 | 13,072 | 410 | 7,799 | 28.5 (18.0 to 37.8) |
| 5<sup>th</sup> month after Dose 2 and no Dose 3 | 622 | 12,981 | 403 | 7,811 | 10.6 (-2.3 to 21.9) |
| 6<sup>th</sup> month after Dose 2 and no Dose 3 | 1,510 | 13,087 | 987 | 7,816 | 14.3 (6.2 to 21.8) |
| 7<sup>th</sup> month after Dose 2 and no Dose 3 | 2,425 | 13,238 | 1,529 | 7,894 | 9.6 (2.4 to 16.3) |
| 8<sup>th</sup> month after Dose 2 and no Dose 3 | 3,625 | 13,026 | 2,003 | 7,915 | -7.5 (-15.3 to -0.2) |
| 9<sup>th</sup> month after Dose 2 and no Dose 3 | 2,989 | 13,134 | 1,779 | 7,901 | 1.5 (-6.2 to 8.7) |
| 10<sup>th</sup> month after Dose 2 and no Dose 3 | 6,257 | 12,946 | 3,340 | 7,974 | -17.7 (-25.6 to -10.3) |
| 11<sup>th</sup> month after Dose 2 and no Dose 3 | 2,335 | 13,045 | 1,337 | 7,867 | -0.3 (-10.2 to 8.6) |
| ≥12<sup>th</sup> month after Dose 2 and no Dose 3 | 614 | 12,984 | 415 | 7,811 | 16.5 (3.1 to 28.1) |
| **Dose 3 (booster dose)** |  |  |  |  |  |
| 1 week after Dose 3 | 403 | 12,945 | 281 | 7,800 | 15.8 (0.9 to 28.4) |
| 2-3 weeks after Dose 3 | 603 | 13,238 | 703 | 7,775 | 53.6 (47.4 to 59.1) |
| 4-5 weeks after Dose 3 | 666 | 13,304 | 771 | 7,772 | 56.6 (50.8 to 61.7) |
| 6-7 weeks after Dose 3 | 949 | 13,325 | 888 | 7,793 | 46.2 (39.7 to 52.0) |
| 8-9 weeks after Dose 3 | 521 | 13,082 | 470 | 7,797 | 38.0 (28.1 to 46.5) |
| 10-11 weeks after Dose 3 | 334 | 13,068 | 328 | 7,816 | 43.7 (32.9 to 52.7) |
| ≥12 weeks after Dose 3 | 719 | 13,139 | 618 | 7,811 | 37.6 (28.8 to 45.4) |
| **mRNA-1273** |  |  |  |  |  |
| **Dose 1** |  |  |  |  |  |
| 0-13 days after Dose 1 | 19 | 12,761 | 10 | 7,709 | -13.6 (-146.4 to 47.6) |
| ≥14 days after Dose 1 and no Dose 2 | 58 | 12,763 | 36 | 7,707 | -1.6 (-56.8 to 34.1) |
| **Dose 2** |  |  |  |  |  |
| 1-3 months after Dose 2 and no Dose 3 | 44 | 12,797 | 46 | 7,708 | 44.8 (16.0 to 63.8) |
| 4-6 months after Dose 2 and no Dose 3 | 1,924 | 13,137 | 1,363 | 7,755 | 20.8 (13.7 to 27.4) |
| ≥7 months after Dose 2 and no Dose 3 | 6,243 | 12,980 | 3,477 | 7,922 | -9.3 (-16.3 to -2.8) |
| **Dose 3 (booster dose)** |  |  |  |  |  |
| 1 week after Dose 3 | 114 | 12,770 | 70 | 7,708 | 3.6 (-31.0 to 29.1) |
| 2-3 weeks after Dose 3 | 149 | 12,873 | 182 | 7,708 | 53.1 (40.7 to 62.8) |
| 4-5 weeks after Dose 3 | 117 | 12,861 | 146 | 7,699 | 54.6 (41.1 to 65.0) |
| ≥6 weeks after Dose 3 | 117 | 12,828 | 110 | 7,712 | 38.6 (19.4 to 53.1) |

Abbreviations: CI, confidence interval; PCR, polymerase chain reaction.
\*A symptomatic infection was defined as a PCR-positive nasopharyngeal swab conducted because of clinical suspicion due to presence of symptoms compatible with a respiratory tract infection.
<sup>†</sup>In each analysis for a specific time-since-vaccination stratum, we included only those vaccinated in this specific time-since-vaccination stratum and those unvaccinated. Only matched pairs of PCR-positive and PCR-negative persons, in which both members of the pair were either unvaccinated or fell within each time-since-vaccination stratum have been included in the corresponding vaccine effectiveness estimate. Thus, the number of cases (and controls) varied across time-since-vaccination analyses.
<sup>‡</sup>Cases and controls were matched two-to-one by sex, 10-year age group, nationality, and calendar week of PCR test.
<sup>§</sup>Vaccine effectiveness was estimated using the test-negative, case-control study design.<sup>8,9</sup>

**Table 2.** Effectiveness of the BNT162b2 and mRNA-1273 vaccines against any severe, critical, or fatal COVID-19.

| Sub-studies<sup>*</sup> | BNT162b2 |  |  |  |  | mRNA-1273 |  |  |  |  |
| --- | --- | --- | --- | --- | --- | --- | --- | --- | --- | --- |
|  | Cases<sup>†</sup><br>(Severe, critical, or<br>fatal disease)<sup>‡</sup> |  | Controls<sup>†</sup><br>(PCR-negative) |  | Effectiveness in %<br>(95% CI)<sup>§</sup> | Cases<sup>†</sup><br>(Severe, critical, or<br>fatal disease)<sup>‡</sup> |  | Controls<sup>†</sup><br>(PCR-negative) |  | Effectiveness in %<br>(95% CI)<sup>§</sup> |
|  | Vaccinated |  | Vaccinated |  |  | Vaccinated |  | Vaccinated |  |  |
|  | Yes | No | Yes | No |  | Yes | No | Yes | No |  |
| **Dose 1** |  |  |  |  |  |  |  |  |  |  |
| Dose 1 and no Dose 2 | 2 | 105 | 10 | 280 | 46.8 (-1.6 to 89.2) | 0 | 103 | 2 | 280 | 100.0 (Omitted)<sup>¶</sup> |
| **Dose 2** |  |  |  |  |  |  |  |  |  |  |
| 1-6 months after Dose 2 and no Dose 3 | 10 | 112 | 99 | 254 | 73.7 (46.8 to 87.0) | 3 | 105 | 35 | 265 | 76.9 (19.2 to 93.4) |
| ≥7 months after Dose 2 and no Dose 3 | 71 | 134 | 445 | 209 | 80.7 (71.3 to 87.0) | 23 | 117 | 139 | 257 | 64.0 (39.1 to 78.7) |
| **Dose 3 (booster dose)** |  |  |  |  |  |  |  |  |  |  |
| 1-6 weeks after Dose 3 | 7 | 115 | 140 | 238 | 90.6 (77.8 to 96.0) | 1 | 103 | 19 | 270 | 80.8 (-51.9 to 97.6) |
| ≥7 weeks after Dose 3 | 12 | 125 | 199 | 230 | 90.8 (81.5 to 95.5) | 0 | 102 | 5 | 278 | 100.0 (Omitted)<sup>¶</sup> |

Abbreviations: CI, confidence interval; PCR, polymerase chain reaction.
<sup>\*</sup>In each analysis for a specific time-since-vaccination stratum, we included only those vaccinated in this specific time-since-vaccination stratum and those unvaccinated. Only matched pairs of PCR-positive and PCR-negative persons, in which both members of the pair were either unvaccinated or fell within each time-since-vaccination stratum have been included in the corresponding vaccine effectiveness estimate. Thus, the number of cases (and controls) varied across time-since-vaccination analyses.
<sup>†</sup>Cases and controls were matched one-to-five by sex, 10-year age group, nationality, and calendar week of PCR test.
<sup>‡</sup>Severity,<sup>21</sup> criticality,<sup>21</sup> and fatality<sup>22</sup> were defined as per World Health Organization guidelines.
<sup>§</sup>Vaccine effectiveness was estimated using the test-negative, case-control study design.<sup>8,9</sup>
<sup>¶</sup>Confidence interval could not be estimated using conditional logistic regression because of zero events among those vaccinated.

mRNA-1273 effectiveness against symptomatic Omicron infection was highest at 44.8% (95% CI: 16.0-63.8%) in the first three months after the second dose, before gradually declining to negligible levels thereafter (Figure 1 and Table 1). After the booster, effectiveness rapidly rebounded to peak at about 55% between 2-5 weeks after the booster, but then declined again thereafter. Effectiveness against severe, critical, or fatal COVID-19 was high at >60% after the second dose and at >80% after the booster, but the confidence intervals were wide owing to the small number of cases (Table 2).

## Discussion and conclusions

BNT162b2 and mRNA-1273 vaccines show a similar level and pattern of protection against symptomatic Omicron infection. Protection against Omicron is lower than that against Alpha, Beta, and Delta variants,^4,5,14^ and wanes more rapidly than against earlier variants after the second and booster doses.^4,5^ However, the waning appears slower after the booster compared to after the second dose. Meanwhile, protection against hospitalization and death appears robust and durable after both the second and booster doses.

### Caveats and limitations

Since the immunization campaigns prioritized vaccination of persons with severe or multiple chronic conditions and by age group, the observed pattern of waning of protection could theoretically be confounded by effects of age and comorbidities. Individual-level data on co-morbid conditions were not available; therefore, they could not be explicitly factored into our analysis. However, only a small proportion of the study population may have had serious co-morbid conditions. Only 9% of the population of Qatar are ≥50 years of age,^6,7^ and 60% are young, expatriate craft and manual workers working in mega-development projects.^19,20,27^ The national list of persons prioritized to receive the vaccine during the first phase of vaccine roll-out included only 19,800 individuals of all age groups with serious co-morbid conditions. Old age may serve as a partial proxy for co-morbid conditions. A similar pattern of waning of protection was observed for younger and older persons in our earlier waning of vaccine effectiveness studies for the same population.^4,5^ However, with the small proportion of Qatar’s population being ≥50 years of age,^6,7^ our findings may not be generalizable to other countries in which elderly citizens constitute a larger proportion of the total population.

Vaccinated persons presumably have a higher social contact rate than unvaccinated persons, and they may also adhere less strictly to safety measures.^28-30^ This behavior could reduce real-world effectiveness of the vaccine compared to its biological effectiveness, possibly explaining the waning of protection. Public health restrictions in Qatar were eased gradually, but differently for vaccinated and unvaccinated persons. Many social, work, and travel activities presently require evidence of vaccination (a “health pass”) that is administered through a mandatory mobile app (the Ehteraz app). However, risk compensation is unlikely to solely explain the observed waning of protection over time.

PCR testing in Qatar is done on a mass scale, such that about 5% of the population are tested every week.^4^ With the large demand for testing during the Omicron wave, use of rapid antigen testing (RAT) was expanded to supplement PCR testing at health care facilities starting from January 5, 2022, but information on reason for testing was not available for the RAT testing to be included in our study.

Effectiveness was assessed using an observational, test-negative, case-control study design,^8,9^ rather than a design in which cohorts of vaccinated and unvaccinated individuals were followed up. However, the cohort study design applied earlier to the same population of Qatar yielded findings similar to those of the test-negative case-control design,^13-16^ supporting the validity of this standard approach in assessing vaccine effectiveness for respiratory tract infections.^8-16^ Moreover, our recent study of the effectiveness of booster vaccination against SARS-CoV-2 symptomatic infection, relative to that of the primary series, used a cohort study design^3^ and its results are consistent with the results generated in the present study using the test-negative, case-control study design.

With the high COVID-19 vaccine coverage in Qatar (>80%), the majority of those unvaccinated were children or young persons, and thus the reference group of those unvaccinated is not representative of the total population demographics. However, matching by age was implemented in all analyses to minimize potential bias arising from the young population structure of those unvaccinated.

To rapidly scale up vaccination, some vaccination campaigns are conducted outside healthcare facilities; thus, records of vaccination are not immediately uploaded into the CERNER system, which tracks all vaccination records at the national level. This administrative time delay can introduce a misclassification bias of those vaccinated versus those unvaccinated. A sensitivity analysis investigating the impact of such potential bias, by assuming a 10% misclassification bias of those vaccinated, found a difference of, at most, few percentage points in estimated effectiveness.^5^ A key strength of the test-negative, case-control study design is that it is less susceptible to this form of bias.^8,9^

Nonetheless, one cannot exclude the possibility that in real-world data, bias could arise in unexpected ways, or from unknown sources, such as subtle differences in test-seeking behavior or changes in the pattern of testing with introduction of other testing modalities, such as RAT. For example, with the large Omicron wave in Qatar, use of RAT was expanded to supplement PCR testing starting from January 5, 2022. However, RAT was broadly implemented in the population and thus may not have differentially affected PCR testing to introduce bias.

With the small number of PCR-positive symptomatic infections in some of the time-since-vaccination strata for those vaccinated with mRNA-1273, it was not possible to estimate mRNA-1273 effectiveness month-by-month after the second dose. Furthermore, with the relatively small number of cases and controls that progressed to severe, critical, or fatal COVID-19 during the Omicron wave, confidence intervals for vaccine effectiveness against hospitalization and death were either very wide or could not be estimated, especially so for the mRNA-1273 analyses.

Notwithstanding these limitations, consistent findings were reached indicating rapid waning of vaccine protection against symptomatic Omicron infection that are broadly consistent with findings of other studies.^31-39^ Moreover, with the mass scale of PCR testing in Qatar,^4^ the likelihood of bias is perhaps minimized. Extensive sensitivity and additional analyses were conducted to investigate effects of potential bias in our earlier studies for the BNT162b2^4^ and mRNA-1273^5^ vaccines, which used the same methodology as the present study. These included different adjustments in the analysis, different approaches for factoring prior infection in the analysis, and different study inclusion and exclusion criteria to investigate whether the effectiveness estimates could have been biased.^4,5^ All analyses showed consistent findings.^4,5^

## Data Availability

The dataset of this study is a property of the Qatar Ministry of Public Health that was provided to the researchers through a restricted-access agreement that prevents sharing the dataset with a third party or publicly. Future access to this dataset can be considered through a direct application for data access to Her Excellency the Minister of Public Health (https://www.moph.gov.qa/english/Pages/default.aspx). Aggregate data are available within the manuscript and its Supplementary information.

## Acknowledgements

We acknowledge the many dedicated individuals at Hamad Medical Corporation, the Ministry of Public Health, the Primary Health Care Corporation, the Qatar Biobank, Sidra Medicine, and Weill Cornell Medicine – Qatar for their diligent efforts and contributions to make this study possible.

The authors are grateful for support from the Biomedical Research Program and the Biostatistics, Epidemiology, and Biomathematics Research Core, both at Weill Cornell Medicine-Qatar, as well as for support provided by the Ministry of Public Health, Hamad Medical Corporation, and Sidra Medicine. The authors are also grateful for the Qatar Genome Programme and Qatar University Biomedical Research Center for institutional support for the reagents needed for the viral genome sequencing. Statements made herein are solely the responsibility of the authors. The funders of the study had no role in study design, data collection, data analysis, data interpretation, or writing of the article.

## Author contributions

HC co-designed the study, performed the statistical analyses, and co-wrote the first draft of the article. LJA conceived and co-designed the study, led the statistical analyses, and co-wrote the first draft of the article. PT and MRH conducted the multiplex, RT-qPCR variant screening and viral genome sequencing. HY, HAK, and MKS conducted viral genome sequencing. All authors contributed to data collection and acquisition, database development, discussion and interpretation of the results, and to the writing of the manuscript. All authors have read and approved the final manuscript.

## Supplementary Appendix

### Section S1. COVID-19 severity, criticality, and fatality classification

Severe Coronavirus Disease 2019 (COVID-19) disease was defined per the World Health Organization (WHO) classification as a severe acute respiratory syndrome coronavirus 2 (SARS-CoV-2) infected person with “oxygen saturation of <90% on room air, and/or respiratory rate of >30 breaths/minute in adults and children >5 years old (or ≥60 breaths/minute in children <2 months old or ≥50 breaths/minute in children 2-11 months old or ≥40 breaths/minute in children 1–5 years old), and/or signs of severe respiratory distress (accessory muscle use and inability to complete full sentences, and, in children, very severe chest wall indrawing, grunting, central cyanosis, or presence of any other general danger signs)”.^1^ Detailed WHO criteria for classifying SARS-CoV-2 infection severity can be found in the WHO technical report.^1^

Critical COVID-19 disease was defined per WHO classification as a SARS-CoV-2 infected person with “acute respiratory distress syndrome, sepsis, septic shock, or other conditions that would normally require the provision of life sustaining therapies such as mechanical ventilation (invasive or non-invasive) or vasopressor therapy”.^1^ Detailed WHO criteria for classifying SARS-CoV-2 infection criticality can be found in the WHO technical report.^1^

COVID-19 death was defined per WHO classification as “a death resulting from a clinically compatible illness, in a probable or confirmed COVID-19 case, unless there is a clear alternative cause of death that cannot be related to COVID-19 disease (e.g. trauma). There should be no period of complete recovery from COVID-19 between illness and death. A death due to COVID-19 may not be attributed to another disease (e.g. cancer) and should be counted independently of preexisting conditions that are suspected of triggering a severe course of COVID-19”. Detailed WHO criteria for classifying COVID-19 death can be found in the WHO technical report.^2^

### Section S2. Laboratory methods and variant ascertainment

#### Real-time reverse-transcription polymerase chain reaction testing

Nasopharyngeal and/or oropharyngeal swabs were collected for PCR testing and placed in Universal Transport Medium (UTM). Aliquots of UTM were: extracted on a QIAsymphony platform (QIAGEN, USA) and tested with real-time reverse-transcription PCR (RT-qPCR) using TaqPath COVID-19 Combo Kits (Thermo Fisher Scientific, USA) on an ABI 7500 FAST (Thermo Fisher, USA); tested directly on the Cepheid GeneXpert system using the Xpert Xpress SARS-CoV-2 (Cepheid, USA); or loaded directly into a Roche cobas 6800 system and assayed with a cobas SARS-CoV-2 Test (Roche, Switzerland). The first assay targets the viral S, N, and ORF1ab gene regions. The second targets the viral N and E-gene regions, and the third targets the ORF1ab and E-gene regions.

All PCR testing was conducted at the Hamad Medical Corporation Central Laboratory or Sidra Medicine Laboratory, following standardized protocols.

#### Classification of infections by variant type

Surveillance for SARS-CoV-2 variants in Qatar is mainly based on viral genome sequencing and multiplex RT-qPCR variant screening^3^ of random positive clinical samples,^4-9^ complemented by deep sequencing of wastewater samples.^6,10^

A total of 315 random SARS-CoV-2-positive specimens collected between December 19, 2021 and January 22, 2022 were viral whole-genome sequenced on a Nanopore GridION sequencing device. Of these, 300 (95.2%) were confirmed as Omicron infections and 15 (4.8%) as Delta (B.1.617.2)^11^ infections.^6,12^ No Delta case was detected in the viral genome sequencing since January 8, 2022.

Additionally, a total of 1,315 random SARS-CoV-2-positive specimens collected between December 22, 2021 and January 1, 2022 were RT-qPCR genotyped. The RT-qPCR genotyping identified 1 B.1.617.2-like Delta case, 366 BA.1-like Omicron cases, 898 BA.2-like Omicron cases, and 50 were undetermined cases where the genotype could not be assigned.

The accuracy of the RT-qPCR genotyping was verified against either Sanger sequencing of the receptor-binding domain (RBD) of SARS-CoV-2 surface glycoprotein (S) gene, or by viral whole-genome sequencing on a Nanopore GridION sequencing device. From 147 random SARS-CoV-2-positive specimens all collected in December of 2021, RT-qPCR genotyping was able to assign a genotype in 129 samples. The agreement between RT-qPCR genotyping and sequencing was 100% for Delta (n=82), 100% for Omicron BA.1 (n=18), and 93% for Omicron BA.2 (27 of 29 were correctly assigned to BA.2 and remaining 2 specimens genotyped as BA.2 were B.1.617.2 by sequencing). Of the remaining 18 specimens: 10 failed PCR amplification and sequencing, 8 could not be assigned a genotype by RT-qPCR (4 of 8 were B.1.617.2 by sequencing, and the remaining 4 failed sequencing). All the variant RT-qPCR genotyping was conducted at the Sidra Medicine Laboratory following standardized protocols.

The large Omicron-wave exponential-growth phase in Qatar started on December 19, 2021 and peaked in mid-January, 2022.^6,12,13^ The study duration coincided with the intense Omicron wave where Delta incidence was limited and contributed less than 10% of cases. Accordingly, any PCR-positive test during the study duration, between December 23, 2021 and February 2, 2022, was used as a proxy for Omicron infection. Of note that the study duration started on December 23, 2021, and not on December 19, 2021, to minimize the occurrence of residual Delta incidence during the first few days of the Omicron wave.

**Table S1.** STROBE checklist for case-control studies.

| Table S1: Checklist for case-control studies |  |  |  |
| --- | --- | --- | --- |
|  | Item No | Recommendation | Main text page |
| Title and abstract | 1 | (a) Indicate the study's design with a commonly used term in the title or the abstract | Abstract |
|  |  | (b) Provide in the abstract an informative and balanced summary of what was done and what was found | Abstract |
| Introduction |  |  |  |
| Background/rationale | 2 | Explain the scientific background and rationale for the investigation being reported | Introduction |
| Objectives | 3 | State specific objectives, including any prespecified hypotheses | Introduction |
| Methods |  |  |  |
| Study design | 4 | Present key elements of study design | Methods ('Study population, data sources, and study design') |
| Setting | 5 | Describe the setting, locations, and relevant dates, including periods of recruitment, exposure, follow-up, and data collection | Methods ('Study population, data sources, and study design') & Supp. Figure S1 |
| Participants | 6 | (a) Give the eligibility criteria, and the sources and methods of case ascertainment and control selection. Give the rationale for the choice of cases and controls<br>(b) For matched studies, give matching criteria and the number of controls per case | Methods ('Study population, data sources, and study design') & Supp. Figure S1 |
| Variables | 7 | Clearly define all outcomes, exposures, predictors, potential confounders, and effect modifiers. Give diagnostic criteria, if applicable | Methods ('Study population, data sources, and study design' & 'Statistical analysis'), Supp. Sections S1 & S2, & Supp. Figure S1 |
| Data sources/measurement | 8 | For each variable of interest, give sources of data and details of methods of assessment (measurement). Describe comparability of assessment methods if there is more than one group | Methods ('Study population, data sources, and study design' & 'Statistical analysis') & Supp. Table S2 |
| Bias | 9 | Describe any efforts to address potential sources of bias | Methods ('Study population, data sources, and study design' & 'Statistical analysis') |
| Study size | 10 | Explain how the study size was arrived at | Supp. Figure S1 |
| Quantitative variables | 11 | Explain how quantitative variables were handled in the analyses. If applicable, describe which groupings were chosen and why | Methods ('Study population, data sources, and study design' & 'Statistical analysis') & Supp. Table S2 |
| Statistical methods | 12 | (a) Describe all statistical methods, including those used to control for confounding | Methods ('Statistical analysis') |
|  |  | (b) Describe any methods used to examine subgroups and interactions | Methods ('Statistical analysis') |
|  |  | (c) Explain how missing data were addressed | NA, see Methods ('Study population, data sources, and study design') |
|  |  | (d) If applicable, explain how matching of cases and controls was addressed | Methods ('Study population, data sources, and study design' & 'Statistical analysis') |
|  |  | (e) Describe any sensitivity analyses | NA, see Discussion ('Caveats and limitations') |
| Results |  |  |  |
| Participants | 13 | (a) Report numbers of individuals at each stage of study—eg numbers potentially eligible, examined for eligibility, confirmed eligible, included in the study, completing follow-up, and analysed<br>(b) Give reasons for non-participation at each stage<br>(c) Consider use of a flow diagram | Supp. Figure S1 |
| Descriptive data | 14 | (a) Give characteristics of study participants (eg demographic, clinical, social) and information on exposures and potential confounders<br>(b) Indicate number of participants with missing data for each variable of interest | Supp. Tables S2-S3<br>NA, see Methods ('Study population, data sources, and study design') |
| Outcome data | 15 | Report numbers in each exposure category, or summary measures of exposure | Results, Figure 1, & Tables 1-2 |
| Main results | 16 | (a) Give unadjusted estimates and, if applicable, confounder-adjusted estimates and their precision (eg, 95% confidence interval). Make clear which confounders were adjusted for and why they were included | Results, Figure 1, & Tables 1-2 |
|  |  | (b) Report category boundaries when continuous variables were categorized | Supp. Table S2 |
|  |  | (c) If relevant, consider translating estimates of relative risk into absolute risk for a meaningful time period | NA |
| Other analyses | 17 | Report other analyses done—eg analyses of subgroups and interactions, and sensitivity analyses | NA, see Discussion ('Caveats and limitations') |
| **Discussion** |  |  |  |
| Key results | 18 | Summarise key results with reference to study objectives | Discussion, paragraph 1 |
| Limitations | 19 | Discuss limitations of the study, taking into account sources of potential bias or imprecision. Discuss both direction and magnitude of any potential bias | Discussion ('Caveats and limitations') |
| Interpretation | 20 | Give a cautious overall interpretation of results considering objectives, limitations, multiplicity of analyses, results from similar studies, and other relevant evidence | Discussion, paragraph 1 |
| Generalisability | 21 | Discuss the generalisability (external validity) of the study results | Discussion ('Caveats and limitations') & Supp. Table S3 |
| **Other information** |  |  |  |
| Funding | 22 | Give the source of funding and the role of the funders for the present study and, if applicable, for the original study on which the present article is based | Acknowledgements |

Abbreviations: NA, not applicable; Supp. Supplementary Appendix.

**Figure S1.**
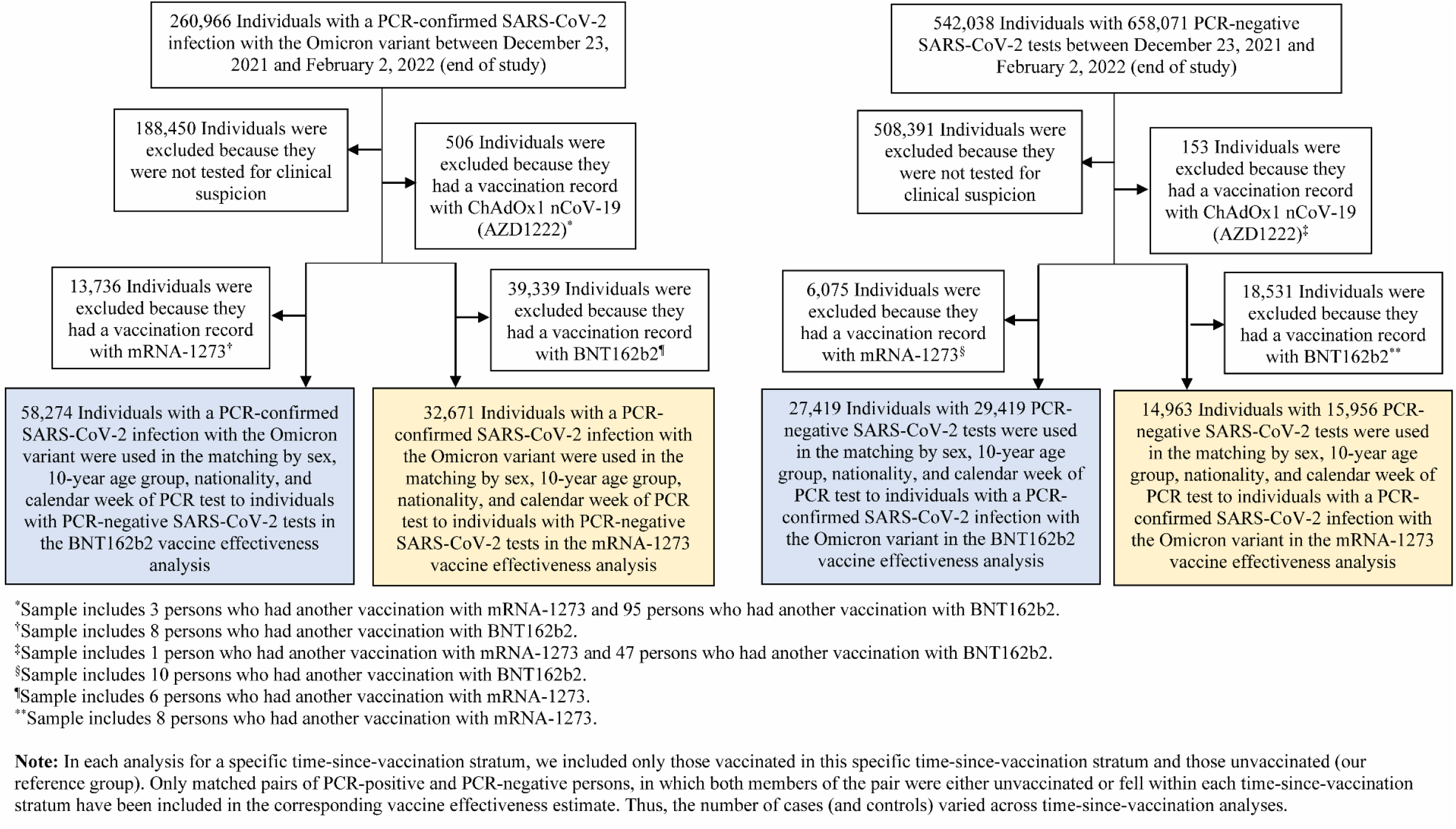
Flowchart describing the population selection process for investigating the effectiveness of the BNT162b2 and mRNA-1273 vaccines during the SARS-CoV-2 Omicron infection wave.

**Table S2.** Demographic characteristics of cases and controls in samples used to estimate the effectiveness of BNT162b2 and mRNA-1273 vaccines against symptomatic SARS-CoV-2 Omicron infection. The table includes samples used in the 1-month-after-second-dose analysis and 4-5-weeks-after-third-dose analysis for BNT162b2, and samples used in the 1-3-months-after-second-dose analysis and 4-5-weeks-after-third-dose analysis for mRNA-1273. These samples provide examples of the study population. Remaining samples for the different time-since-vaccination strata are similar to the table samples.

| Characteristics | BNT162b2 |  |  |  |  |  | mRNA-1273 |  |  |  |  |  |
| --- | --- | --- | --- | --- | --- | --- | --- | --- | --- | --- | --- | --- |
|  | 1-month-after-second-dose |  |  | 4-5-weeks-after-third-dose |  |  | 1-3-months-after-second-dose |  |  | 4-5-weeks-after-third-dose |  |  |
|  | Cases*<br>(PCR-positive) | Controls*<br>(PCR-negative) | SMD† | Cases*<br>(PCR-positive) | Controls*<br>(PCR-negative) | SMD† | Cases*<br>(PCR-positive) | Controls*<br>(PCR-negative) | SMD† | Cases*<br>(PCR-positive) | Controls*<br>(PCR-negative) | SMD† |
|  | N=13,042 | N=7,911 |  | N=13,970 | N=8,543 |  | N=12,841 | N=7,754 |  | N=12,978 | N=7,845 |  |
| **Median age (IQR) — years** | 12 (6-32)‡ | 12 (5-32)‡ | 0.01§ | 18 (6-34)‡ | 19 (5-34)‡ | 0.00§ | 12 (6-32)‡ | 12 (5-32)‡ | 0.02§ | 12 (6-32)‡ | 12 (5-32)‡ | 0.01§ |
| **Age group — no. (%)** |  |  |  |  |  |  |  |  |  |  |  |  |
| <20 years | 7,126 (54.6) | 4,324 (54.7) | 0.01 | 7,101 (50.8) | 4,307 (50.4) | 0.01 | 7,079 (55.1) | 4,291 (55.3) | 0.01 | 7,080 (54.6) | 4,292 (54.7) | 0.01 |
| 20-29 years | 2,090 (16.0) | 1,254 (15.9) |  | 2,147 (15.4) | 1,298 (15.2) |  | 2,034 (15.8) | 1,213 (15.6) |  | 2,030 (15.6) | 1,211 (15.4) |  |
| 30-39 years | 2,148 (16.5) | 1,304 (16.5) |  | 2,524 (18.1) | 1,560 (18.3) |  | 2,084 (16.2) | 1,253 (16.2) |  | 2,168 (16.7) | 1,302 (16.6) |  |
| 40-49 years | 870 (6.7) | 534 (6.8) |  | 1,160 (8.3) | 731 (8.6) |  | 856 (6.7) | 523 (6.7) |  | 877 (6.8) | 540 (6.9) |  |
| 50-59 years | 468 (3.6) | 290 (3.7) |  | 644 (4.6) | 401 (4.7) |  | 463 (3.6) | 280 (3.6) |  | 492 (3.8) | 302 (3.9) |  |
| 60-69 years | 205 (1.6) | 126 (1.6) |  | 243 (1.7) | 154 (1.8) |  | 197 (1.5) | 120 (1.6) |  | 203 (1.6) | 124 (1.6) |  |
| 70+ years | 135 (1.0) | 79 (1.0) |  | 151 (1.1) | 92 (1.1) |  | 128 (1.0) | 74 (1.0) |  | 128 (1.0) | 74 (0.9) |  |
| **Sex** |  |  |  |  |  |  |  |  |  |  |  |  |
| Male | 6,827 (52.4) | 4,168 (52.7) | 0.01 | 7,379 (52.8) | 4,552 (53.3) | 0.01 | 6,748 (52.6) | 4,108 (53.0) | 0.01 | 6,853 (52.8) | 4,171 (53.2) | 0.01 |
| Female | 6,215 (47.7) | 3,743 (47.3) |  | 6,591 (47.2) | 3,991 (46.7) |  | 6,093 (47.5) | 3,646 (47.0) |  | 6,125 (47.2) | 3,674 (46.8) |  |
| **Nationality\*\*** |  |  |  |  |  |  |  |  |  |  |  |  |
| Bangladeshi | 242 (1.9) | 181 (2.3) | 0.05 | 242 (1.7) | 179 (2.1) | 0.04 | 241 (1.9) | 172 (2.2) | 0.04 | 245 (1.9) | 172 (2.2) | 0.04 |
| Egyptian | 881 (6.8) | 505 (6.4) |  | 960 (6.9) | 555 (6.5) |  | 868 (6.8) | 497 (6.4) |  | 873 (6.7) | 501 (6.4) |  |
| Filipino | 1,074 (8.2) | 633 (8.0) |  | 1,316 (9.4) | 795 (9.3) |  | 1,046 (8.2) | 607 (7.8) |  | 1,085 (8.4) | 639 (8.2) |  |
| Indian | 1,732 (13.3) | 1,044 (13.2) |  | 2,021 (14.5) | 1,239 (14.5) |  | 1,701 (13.3) | 1,023 (13.2) |  | 1,775 (13.7) | 1,066 (13.6) |  |
| Nepalese | 250 (1.9) | 137 (1.7) |  | 256 (1.8) | 143 (1.7) |  | 251 (2.0) | 137 (1.8) |  | 255 (2.0) | 140 (1.8) |  |
| Pakistani | 550 (4.2) | 322 (4.1) |  | 570 (4.1) | 336 (3.9) |  | 543 (4.2) | 316 (4.1) |  | 545 (4.2) | 316 (4.0) |  |
| Qatari | 4,382 (33.6) | 2,601 (32.9) |  | 4,475 (32.0) | 2,674 (31.3) |  | 4,284 (33.4) | 2,541 (32.8) |  | 4,300 (33.1) | 2,551 (32.5) |  |
| Sri Lankan | 163 (1.3) | 100 (1.3) |  | 170 (1.2) | 104 (1.2) |  | 160 (1.3) | 98 (1.3) |  | 155 (1.2) | 95 (1.2) |  |
| Sudanese | 631 (4.8) | 381 (4.8) |  | 660 (4.7) | 395 (4.6) |  | 630 (4.9) | 378 (4.9) |  | 630 (4.9) | 377 (4.8) |  |
| Other nationalities†† | 3,137 (24.1) | 2,007 (25.4) |  | 3,300 (23.6) | 2,123 (24.9) |  | 3,117 (24.3) | 1,985 (25.6) |  | 3,115 (24.0) | 1,988 (25.3) |  |

Abbreviations: IQR, interquartile range; PCR, polymerase chain reaction; SMD, standardized mean difference.
\*Cases and controls were matched two-to-one by sex, 10-year age group, nationality, and calendar week of PCR test.
†SMD is the difference in the mean of a covariate between groups divided by the pooled standard deviation. An SMD<0.1 indicates adequate matching.
‡With the high vaccine coverage in Qatar (>80%), the majority of those unvaccinated (reference group) were children or young persons, and thus the young median age in this study sample. For the same reason, most of those in the 1-month-after-second-dose stratum in the BNT162b2 analysis and in the 1-3-months-after-second-dose stratum in the mRNA-1273 analysis were children or adolescents as the majority of adults received their second dose months earlier.
§SMD is for the mean difference between groups divided by the pooled standard deviation.
¶With the high vaccine coverage in Qatar (>80%), the majority of those unvaccinated (reference group) were children or young persons, and thus the young median age in this study sample.
\*\*Nationalities were chosen to represent the most populous groups in Qatar.
††These comprise 56 other nationalities in the 1-month-after-second-dose analysis and 56 other nationalities in the 4-5-weeks-after-third-dose analysis for BNT162b2, and 55 other nationalities in the 1-3-months-after-second-dose analysis and 55 other nationalities in the 4-5-weeks-after-third-dose analysis for mRNA-1273.

**Table S3.**
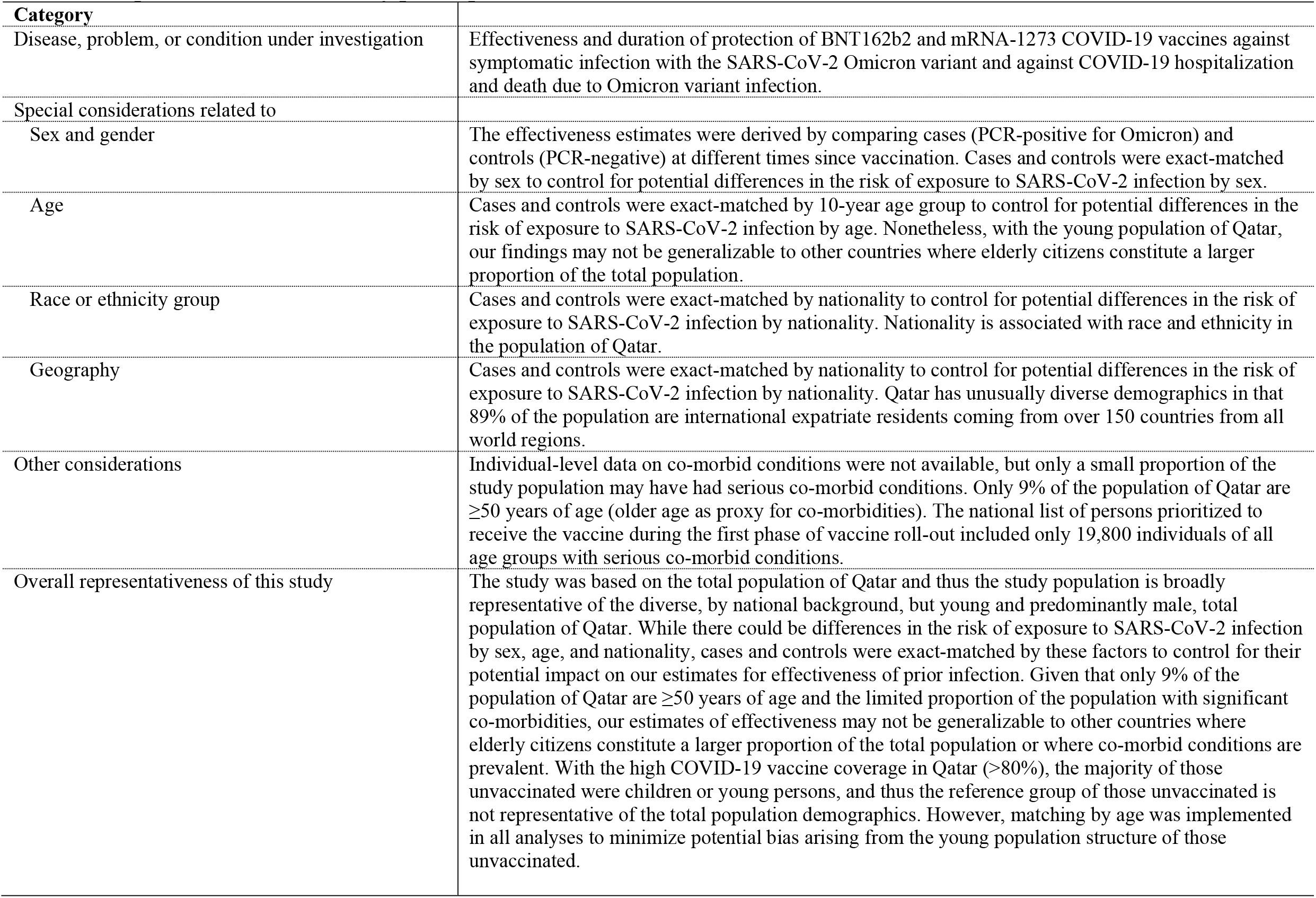
Representativeness of study participants.

